# Screening for amyotrophic lateral sclerosis through interactions with an internet search engine

**DOI:** 10.1101/2023.02.26.23286464

**Authors:** Elad Yom-Tov, Indu Navar, Ernest Fraenkel, James D. Berry

**Affiliations:** Microsoft Research, Israel; EverythingALS, Seattle, Washington, USA; Massachusetts Institute of Technology, Department of Biological Engineering, Cambridge, MA, USA; Sean M. Healey and AMG Center for ALS, Department of Neurology, Massachusetts General Hospital, USA

## Abstract

Amyotrophic lateral sclerosis (ALS), a motor neuron disease, remains a clinical diagnosis with a diagnostic delay of over a year. Here we examine the possibility that interactions with an internet search engine could be used to help screen for ALS. We identified 285 anonymous Bing users whose queries indicated that they had been diagnosed with ALS and matched them to 1) 3276 control users and 2) 1814 users whose searches indicated they had ALS disease mimics. We tested whether the ALS group could be distinguished from controls and disease mimics based on search engine query data. Finally, we conducted a prospective validation from participants who provided access to their Bing search data. The model distinguished between the ALS group and controls with an area under the curve (AUC) of 0.81. Model scores for the ALS group differed from the disease mimics group (ranksum test, P<0.05 with Bonferrini correction). Mild cognitive impairment could not be distinguished from ALS (NS). In the prospective analysis, the model reached an AUC of 0.74. These results suggest that interactions with search engines could be used as a tool to assist in screening for ALS, to reduce diagnostic delay.

## Introduction

Amyotrophic lateral sclerosis (ALS) is an adult-onset neurodegenerative disease that primarily affects the motor neurons in the brain and spinal cord ^1^. The result is a progressive loss of strength that can affect the voluntary muscles of the limbs, bulbar region, and trunk, including muscles of respiration. A small percentage of people with ALS can develop frontotemporal dementia (FTD). A larger percentage of people will develop more subtle cognitive change, particularly affecting executive function and expressive language ^2^.

ALS is typically diagnosed by a neurologist and the diagnosis remains primarily a clinical diagnosis of exclusion shaped by the revised El Escorial Criteria ^3^, though other diagnostic criteria also exist ^4,5^. A medical history and physical examination raise the suspicion of the disease on clinical grounds, and further testing is used to eliminate competing diagnoses and support the clinical impression of the disease ^1^. These additional tests include electromyography to exclude other diseases affecting motor nerves ^4^, imaging and blood tests. The process of diagnosis typically lasts 12 months from symptom onset, and most patients receive at least one incorrect diagnosis during this period ^6,7^. The rate of progression after diagnosis is highly variable between patients, though the duration from symptom onset to diagnosis correlates with rapidity of disease progression, with shorter diagnostic delay correlating with faster progression^8^.

The way that people interact with computers may provide a means to assess cognition ^9^ and could even provide a window into motor function. Motor, language, and executive functions are required to operate the keyboard and mouse, comprehend and create text, plan and focus attention. Changes in search engine queries have been shown to correlate with parts of standard cognitive tests and may thus be used as a continuous, unobtrusive inexpensive monitoring application ^9^. Indeed, internet data, and specifically search engine queries, have been shown to be useful for screening for a variety of diseases including Alzheimer’s Disease ^10^, Parkinson’s Disease ^11^, several types of cancer ^12–14^, diabetes ^15^, and eating disorders ^16^. Similarly, social media postings have been shown to predict depression ^17^, postpartum depression ^18^ and to change prior to emergency department visits ^19^. Therefore, in this study we focus on the question of whether differences in use of and interactions with search engines can be used as a screening tool for ALS.

Past work on screening from internet data used predominantly archival data, where the endpoints used to identify the patient cohort were queries in which users self-identified as having the condition of interest. This offers a method for creating patient cohorts with relatively little recruiting effort, but the patient cohort is not clinically validated, and relying exclusively on this analysis could raise ethical challenges ^20^. An alternative, face-to-face recruitment of patients, is challenging in the case of rare diseases such as ALS.

The current study offers a novel methodology whereby the screening model is developed using archival data of non-clinically verified users. The model is then tested on the data of users who self-identified as having been given an ALS diagnosis by an ALS clinician and provide informed consent to contribute their search engine data anonymously.

## Methods

### Retrospective Study of Existing Search Engine Data

#### Oversight

For this retrospective portion of our study, the methods were performed in accordance with relevant guidelines and regulations and approved by the Microsoft Research institutional review board (IRB).

#### User Identification and Query Characterization

We extracted all queries submitted to the Bing search engine by people in the United States during 2019. Data available for retrospective analysis included the query text, date and time of the query, a pseudo-anonymous user identifier, and data on the interaction between the user and the search results page, specifically, session duration and whether the search engine’s autocorrect function was used. A session is defined as the length of time during which users submit queries to the search engine with inter-query breaks shorter than 30 minutes.

To identify people who are likely to have ALS we found users who queried for phrases that indicated that the user had the disease (referred to as the “reference query”). Specifically, we defined the reference queries to be those containing one of the phrases “I have”, “I have been diagnosed with”, or “I’ve been diagnosed” together with one of the terms “ALS”, “amyotrophic lateral sclerosis”, or “Lou Gehrig’s disease.” We refer to these users as “query-identified” people with ALS.

#### ALS Reference Terms

To identify a control group, we found users who made similar common queries to those of the ALS group. Specifically, we identified the 100,000 most common queries among all users in 2019. We then found, for each user in the ALS group, up to 50 users who made at least 10 of the most common queries and had an overlap of at least 10% with the ALS user.

To identify how frequently query-identified people with ALS and controls searched for key ALS terms, we defined ALS-relevant queries as those including one of the following reference terms:

1. Disease: “ALS”, “amyotrophic lateral sclerosis”, “Lou Gehrig’s disease”
2. Drugs used to treat ALS: “radicava,” “radicut,” “edaravone,” “riluzole,” “rilutek,” “tiglutik,” “nuedexta”
3. Patient groups: “muscular dystrophy association”, “motor neuron disorder association”, “MNDA”, “NEALS”
4. Terms related to the disease: “vital capacity”, “FVC”, “SVC”, “shortness of breath”, “muscle strength”, “muscle weakness”, “twitching”, “fasciculation”, “short breath”.

All query dates of a user were computed relative to the date of the first query relevant to ALS. For users without a history of ALS queries, the reference time was chosen as the middle point between the first and last time that queries from the user were found. Users who queried for fewer than 10 days were excluded.

All queries were characterized by the following attributes: 1) session duration (in seconds); 2) whether the autocorrect was triggered; 3) query length (in characters); 4) number of words in the query; 5) longest word in the query; 6) average query word length.

#### Model Building

A random forest model with 20 trees was trained to distinguish users in the ALS group from those in the control group. Users were then aggregated by the minimal score of their queries (minimum pooling) after removal of the 5 percent of lowest scores per user. Ten-fold cross validation, stratified by users, was applied for estimating model performance. Reference queries were excluded from this analysis.

#### Testing Model Specificity Using Disease Controls

To explore whether the ALS model was specific to the identification of people with ALS, the data used to build the ALS model was used to characterize people with five disease control conditions that have symptoms similar to ALS. These were: 1) Inclusion body myositis, 2) Guillain-Barre syndrome, 3) Cervical radiculopathy, 4) Carpal tunnel syndrome, and 5) Mild cognitive impairment.

Users were identified as disease controls by reference query, as described for ALS above. Or, if too few users made the reference query, they were identified if they queried five or more times about one of the conditions (as in Ofran et al. [13]). Users from these groups who asked about ALS were excluded to avoid contamination of these disease-control groups.

The model developed for ALS was applied to the data from these users and compared to users in the ALS cohort. To reduce the likelihood of overfitting, scores in the ALS group were calculated using 10-fold cross-validation stratified by users. The difference between the scores given to disease control users and the scores given to the ALS users was evaluated using a ranksum test.

### Prospectively Enrolled Study for Model Validation on an External Dataset

As noted, the ALS model was trained on query-identified users. We then conducted an external validation study in people who self-identified as having been diagnosed with ALS by a clinician. These “self-identified” participants enrolled prospectively and contributed existing search engine data for analysis using an automated system.

#### Oversight

For this prospectively enrolled, minimal risk portion of our study, the methods were performed in accordance with relevant guidelines and regulations and approved by the Advarra IRB.

#### Recruitment and Consent

The recruitment and enrollment procedures for the study were designed to select for participants with self-identified ALS diagnoses while preserving anonymity and privacy. Participants were recruited through advertising to members of EverythingALS, a large US-based advocacy organization for people living with ALS. Advertising was IRB approved and was included in webinars, emails to the EverythingALS mailing list and at events. People were asked to direct their browser to the study website where the informed consent was found. Participants read the written informed consent document on a secure webpage. They documented consent by checking an opt-in checkbox to indicate their study consent. The checkbox was only available after scrolling through the written consent document. Study documents were only available after participants had documented consent to the study in this way.

Once consented, participants were asked to provide age and sex and to answer six questions to verify their diagnosis of ALS (Appendix 1). These questions were modelled after those used in the self-registration portion of the US National ALS Registry ^22,23^. Finally, participants were asked to click on a link which would associate their existing search engine data to their study profile. This link created a unique, randomly assigned code recorded in their study record and entered as a query in Bing. The user could then be identified in Bing using the unique code and all other queries associated with the user could be analyzed for this project.

The control group for model validation were all US-based Bing users who were active during 2021, when the study was recruiting self-identified ALS participants, or when they enrolled prospectively and provided data indicating that they had not been diagnosed with ALS. As has been done in other studies, we included controls who might be concerned about ALS, but likely did not have ALS ^24^ because ultimately these are the groups of users it is important to differentiate. We identified control users who queried for ALS one to five times, were active for at least one day and queried at least five times.

## Results

We identified 285 query-identified ALS users and 3276 control users. A total of 485 users in the control group also mentioned reference terms in their queries. However, the ALS group asked, on average, 22.0 queries each that mentioned the ALS-relevant terms compared to controls, who asked 0.6 queries each (ranksum, p<10^−20^).

Users in the ALS group made, on average, 9 queries per day compared to 48 per day by users in the control group (ranksum, p<10^−20^). Users of the ALS group were active for 135 days, on average, compared to 249 days in the controls (ranksum, p<10^−20^).

All six query attributes evaluated were significantly different between the query-identified ALS and control groups (Figure 1a-f). Specifically, people in the ALS group had shorter sessions; they also used longer queries containing longer words and triggered spelling autocorrect more frequently (Table 1).

**Figure 1:**
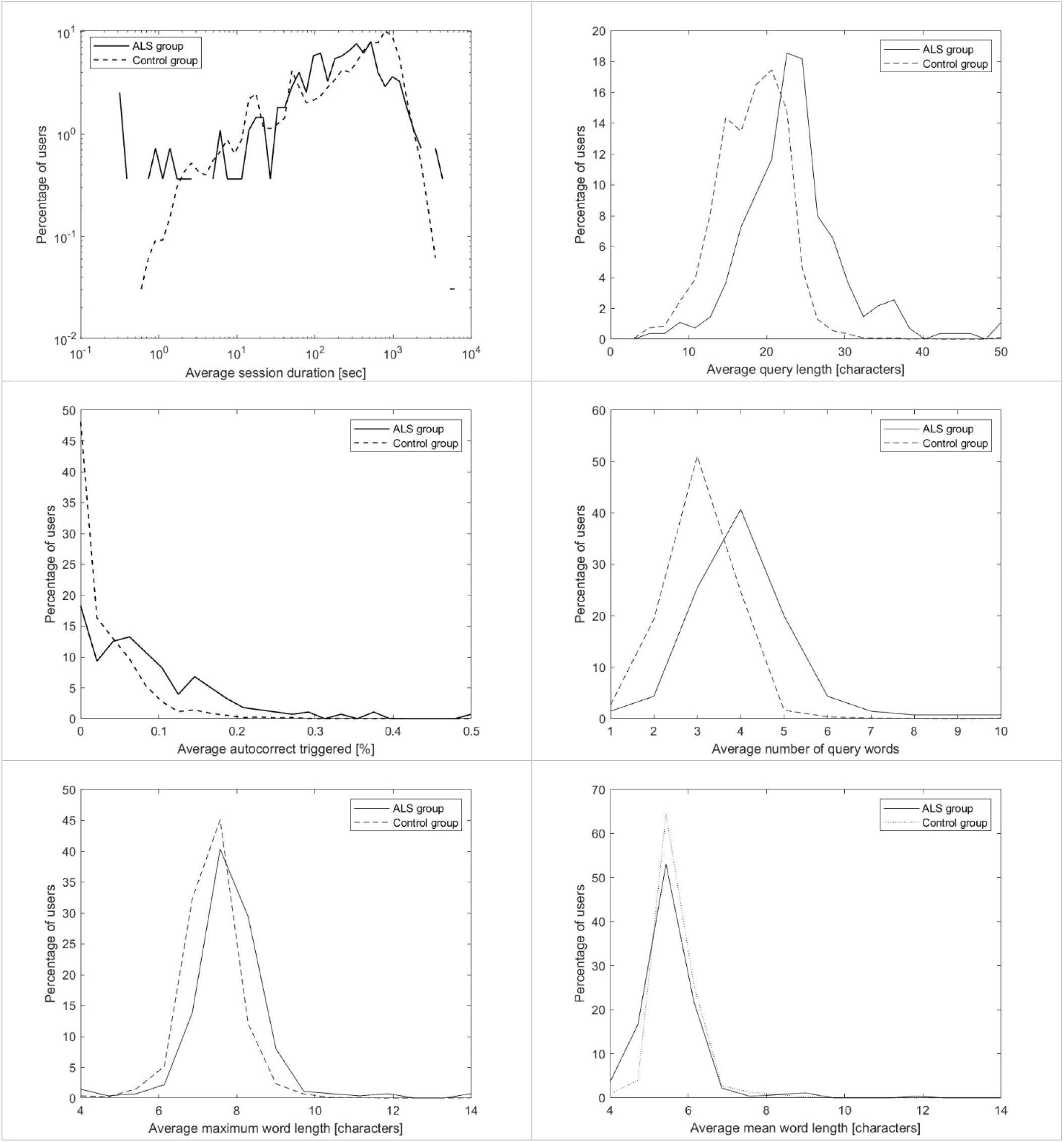
Distribution of query attributes. The distribution of six attributes in the query-identified ALS group and the control group. Average values of each attribute were calculated for each user and the distributions of these average values are shown. Groups differ for all attributes (Wilcoxon sign rank test, P<10^−5^).

**Table 1:** Search Characteristics in ALS and Control Groups.

| Population | Average session duration |  | Average query length |  | Avg autocorrect triggered |  | Avg number of query words |  | Average maximum word length |  | Average mean word length |  |
| --- | --- | --- | --- | --- | --- | --- | --- | --- | --- | --- | --- | --- |
|  | Seconds | p-value | Characters | p-value | Percent | p-value | Words | p-value | Characters | p-value | Characters | p-value |
| ALS | 987.0 |  | 23.6 |  | 0.084 |  | 4.1 |  | 7.8 |  | 5.5 |  |
| Controls | 1104.6 | <0.0001 | 18.1 | <0.0001 | 0.031 | <0.0001 | 3.0 | <0.0001 | 7.4 | <0.0001 | 5.7 | <0.0001 |
| IBM | 868.6 | 0.0155 | 22.4 | 0.0174 | 0.050 | 0.0001 | 3.6 | <0.0001 | 8.0 | 0.0125 | 8.0 | <0.0001 |
| GBS | 751.3 | 0.0207 | 22.3 | 0.2579 | 0.036 | 0.0085 | 3.6 | 0.0348 | 8.0 | 0.4607 | 8.0 | <0.0001 |
| C. Radic | 842.0 | <0.0001 | 22.6 | 0.0015 | 0.069 | 0.0301 | 3.6 | <0.0001 | 8.0 | 0.0033 | 8.0 | <0.0001 |
| CTS | 839.6 | <0.0001 | 22.4 | 0.0015 | 0.070 | 0.1438 | 3.6 | <0.0001 | 8.3 | <0.0001 | 8.3 | <0.0001 |
| MCI | 818.4 | 0.2446 | 25.1 | 0.3563 | 0.069 | 0.5720 | 4.3 | 0.5003 | 8.0 | 0.5877 | 8.0 | <0.0001 |

ALS – amyotrophic lateral sclerosis; IBM – inclusion body myositis; GBS – Guillain-Barre syndrome; C. Radic – cervical radiculopathy; CTS – carpal tunnel syndrome; MCI – mild cognitive impairment

The spelling errors were not attributable to people with ALS asking more about medical terms, leading to more misspelling. While the ALS group queried more about symptoms (2.5% vs. 0.4% in the control population), they made fewer spelling mistakes on symptom queries (8.5%) than on other queries (9.5%). Similarly, people in the ALS population asked more about diseases (6.4% vs 2.6% controls) but had a similar percentage of spelling mistakes on disease queries (9.0%) compared to non-disease-related queries (8.6%). Finally, people in the ALS population asked more about medications (0.8% vs. 0.5% controls) but made fewer spelling mistakes when asking about medications (6.5%) compared to non-medication queries (9.0%).

The final ALS model differentiates query-identified ALS users from controls with an AUC of 0.81 using all data available (Figure 2). For example, this translates to a sensitivity of 74% at a specificity of 74%. Limiting data only within 30 days of the reference query, the AUC is 0.73, and it is similar (0.68) to the AUC when limiting the data to between 90 and 30 days prior to the reference query or to 90 days or more before the reference query (AUC=0.69).

**Figure 2:**
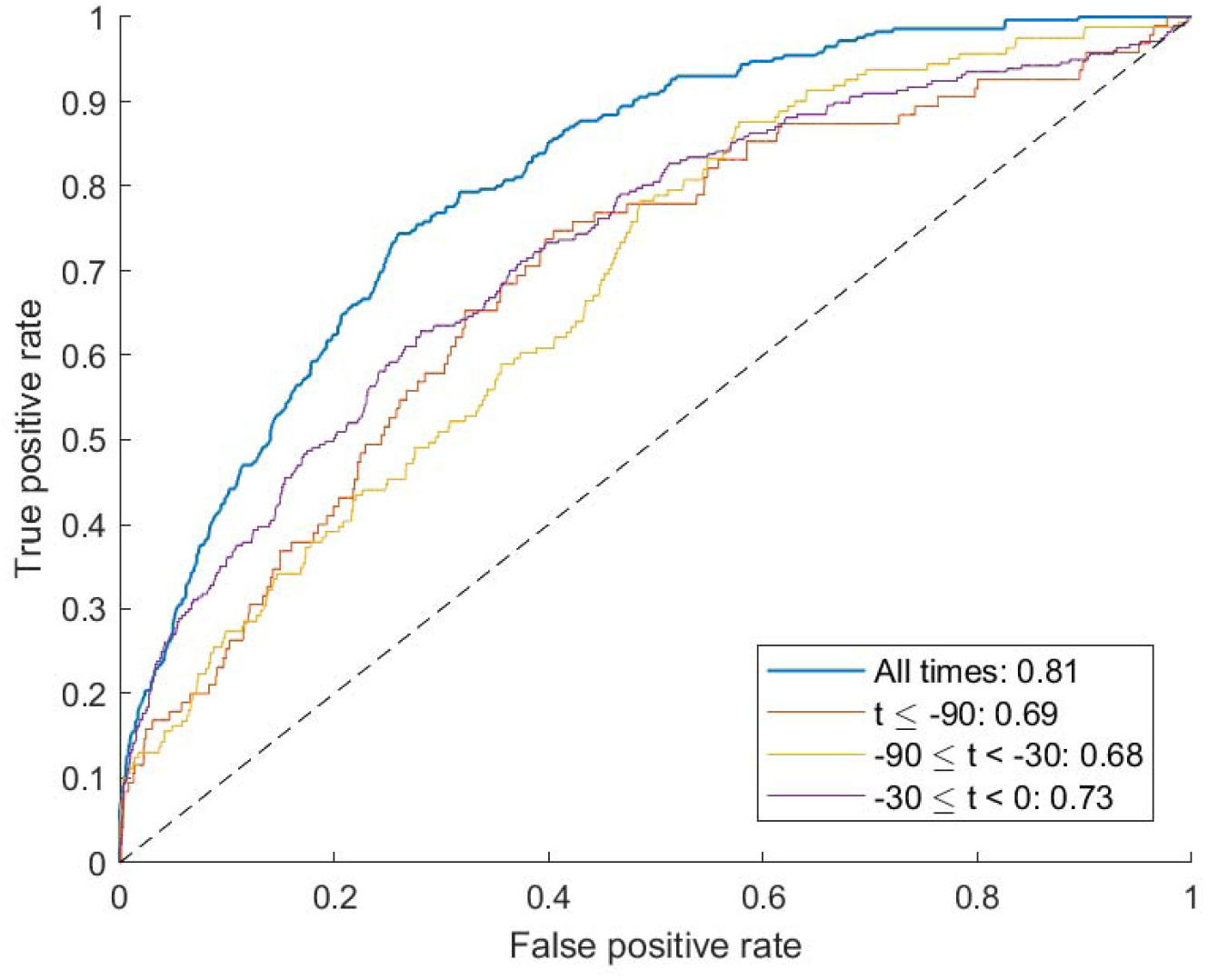
Receiver Operator Curve (retrospective, query-identified users). The area under the curve (AUC) for differentiating query-identified ALS participants from controls was 0.72 using data within 90 days of the reference query and increases to 0.81 using all available data (purple line).

### Evaluating specificity of the ALS model compared to people with related diseases

The distribution of model scores in the ALS cohort was significantly different than for the disease controls Guillain-Barre, inclusion body myositis, cervical radiculopathy, and carpal tunnel syndrome (p<0.05 for all comparisons with Bonferroni correction; Figure 3 and Table 1). The scores for people in the mild cognitive impairment group were not different from those with ALS (NS; Figure 3 and Table 1).

**Figure 3:**
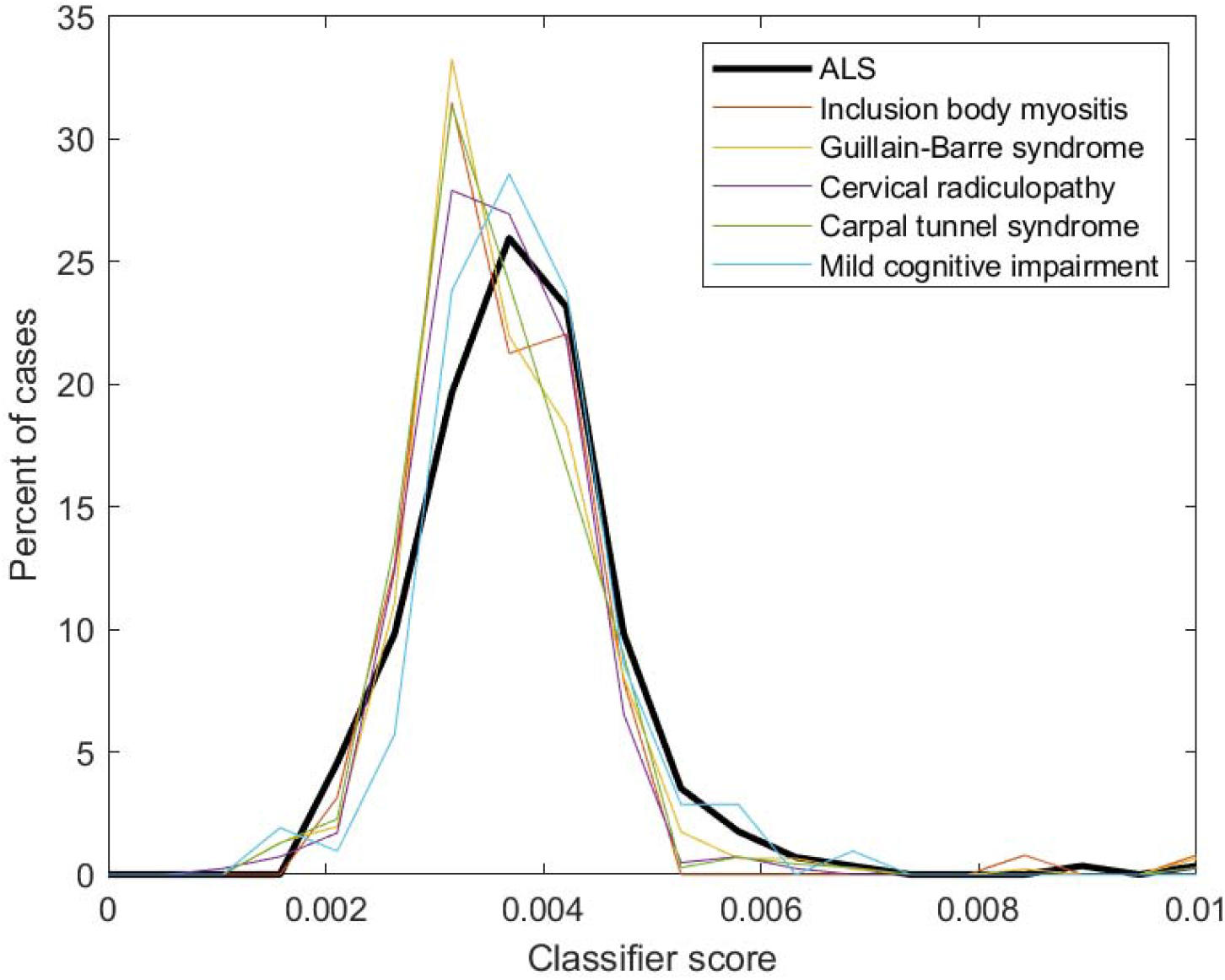
Distribution of the classifier scores. The score distribution for the ALS cohort and for each of the control condition cohorts.

### Model Validation using Prospectively Enrolled External Dataset

For the prospective enrollment of people with self-identified ALS, a total of 122 people visited the study website. Of those, 85 consented to participate in the study and 60 completed the questionnaire. Among consenting participants who completed the questionnaire, 45 reported being diagnosed with ALS or having been told by a health professional that they have or might have ALS. Twelve of those reported they were given an additional diagnosis by a healthcare professional. Data for a total of 21 participants was found in Bing data (35%), of which 16 indicated an ALS diagnosis. Five participants with Bing data indicated that they did not have ALS and were added to the control population. Another 3750 control users were identified, using search engine data as described. Of the consented users with ALS 53% were female, with an average age of 60.2 years (range: 39 – 79, S.D.: 10.3). The median time to participate in the study (among consenting people) was 3.8 minutes. Applying the ALS model to the people with self-identified ALS and controls, the AUC was 0.74 (Figure 4). This translates to a specificity of 86% at 100% sensitivity.

**Figure 4:**
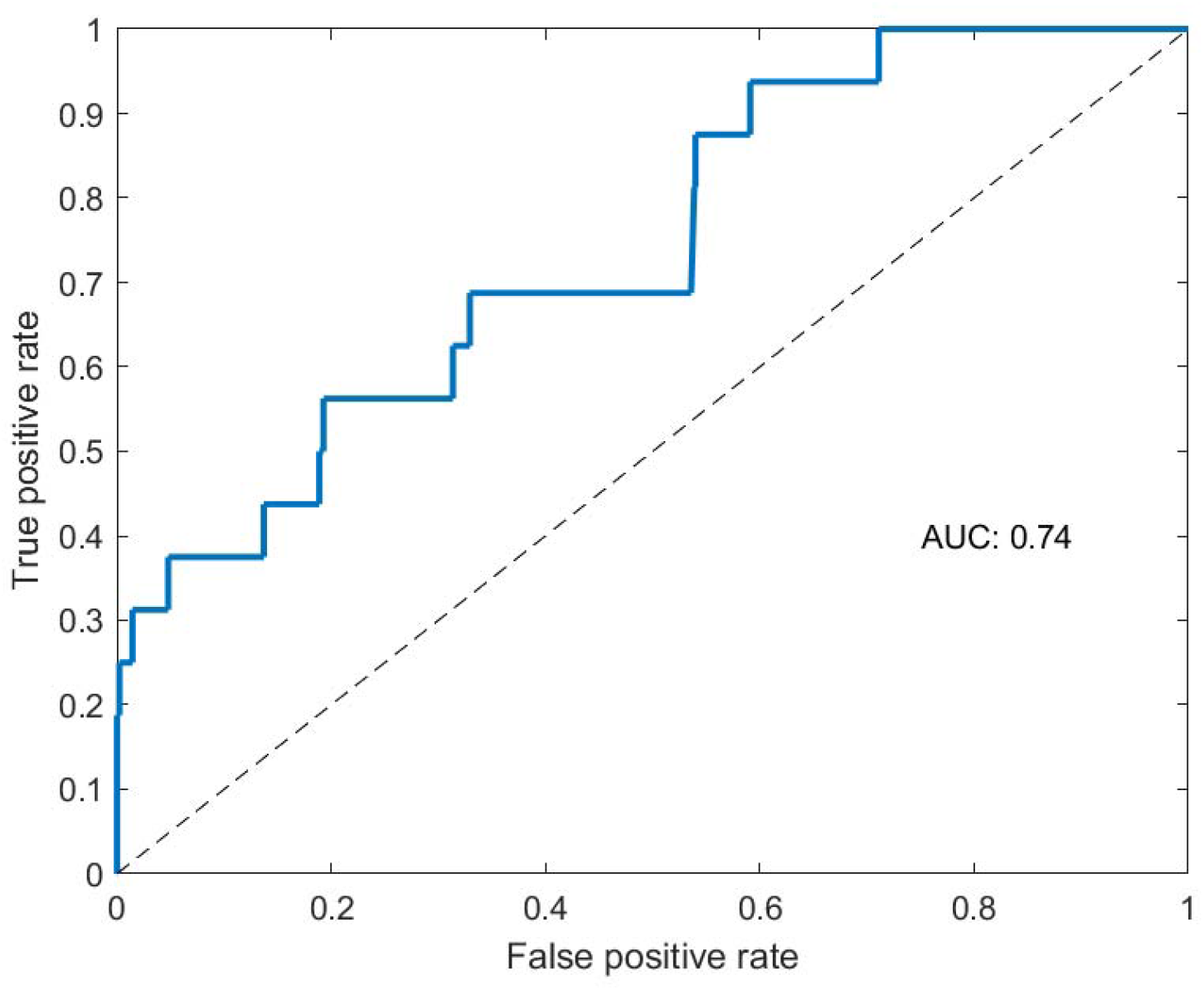
Receiver Operator Curve (prospective, self-identified participants). The AUC distinguishes people with self-identified ALS from controls. The ALS model built on query-identified ALS users and controls (AUC of 0.81 in that population) demonstrates an AUC of 0.73 on this external population with self-identified ALS and controls. In this case, controls were also selected specifically for having searched for ALS-relevant terms 1-5 times.

## Discussion

This study further demonstrates the potential of search engine investigations using data from unidentified users to provide information about the health of populations under-represented in traditional medical research ^20^. Our data suggest that using search engine data, we can recognize a group of users who are likely to have ALS and detect unique search characteristics that distinguish this group from controls with a high specificity and sensitivity. People with ALS queried reference terms more frequently, had fewer queries per day, and queried more uncommon terms. They also triggered the spelling correction significantly more often. Queries from people with ALS may contain more spelling mistakes due to hand weakness causing typographic errors, or, they could have subtle cognitive changes affecting expressive language, which are well documented in ALS and could affect spelling.

One might expect the model performance to change as ALS progresses within a given person. Interestingly, queries at different intervals after study inclusion did not reveal clear changes in the ALS model performance, perhaps because the interval of participation was short and ALS changes were not marked enough over that duration. Thus, these changes may work better as diagnostic markers, rather than markers of ALS progression.

The model was able to distinguish between ALS and disease mimics, except for mild cognitive impairment. Subtle cognitive changes are not uncommon in people with ALS, which may contribute to the challenges distinguishing between these groups. It is interesting that the model was able to distinguish the ALS group from people with conditions that affect motor function, but not cognitive change. Further work is required to test the model’s relative reliance on cognitive changes versus motor changes.

Importantly, the model also performed well on an external validation of a self-identified cohort of people with a diagnosis of ALS.

Our study does have several limitations. First, the data used to train our model was from users identified by their queries. Therefore, we did not have clinical verification for their medical status. Past work has shown that a minority of users indicate an illness when a close family member is the person who is ill and that different demographics query differently, for example, women tend to use longer queries. Thus, queries about illness could be biased by differences in the demographics of the participants ^21^. Furthermore, query-identified people with ALS are certainly only a subset of people with ALS and could represent a biased sample. However, we successfully applied the ALS model to an external population of people with self-reported ALS and found a similar performance, suggesting the model is robust. Second, while we examined behavior within 90 days of the self-identifying query, we are not able to determine the actual time from diagnosis or from first symptoms.

Future work in ALS should be done applying the proposed model to larger cohorts of people with clinically verified ALS to further test its accuracy and to extend the model into different languages. Larger cohorts would also test whether accurate diagnosis can be performed earlier than is currently possible and if the model can help shorten the diagnostic process for ALS.

## Supporting information

Questionnaire

## Data Availability

Due to privacy considerations, data used in this study are confidential. The authors will make data available to researchers upon request, subject to Microsoft Research review and approval.

## Acknowledgements

The authors would like to acknowledge the participants in this study for their effort and dedication to forwarding ALS research.

