## Supplementary material for "Screening for amyotrophic lateral sclerosis through interactions with an internet search engine": Questionnaire

### **Appendix 1: Questionnaire used for participant self-identification of ALS**

1. Were you ever told by a health professional that you might have ALS or Lou Gehrig’s disease?
2. Were you clinically diagnosed with ALS?
3. Have you been seen by a neurologist?
4. What was the date of your diagnosis?
5. Is there another diagnosis that you have been given by a health professional?
6. If so, what was the diagnosis?
